# Long-Term Prognosis and Surgical Outcomes of Culture-Negative Infective Endocarditis

**DOI:** 10.1101/2025.05.28.25328538

**Authors:** Lanlin Zhang, Sheng Yang, Yuanhang Zhai, Yuheng Dong, Guotao Ma, Xinrong Liu, Chaoji Zhang, Shangdong Xu, Jun Zheng

**Affiliations:** Department of Cardiac Surgery, Peking Union Medical College Hospital, Beijing, China

**Keywords:** Infective endocarditis, Culture-negative, Surgery, Prognosis, Risk analysis

## Abstract

**Background:** Culture-negative infective endocarditis (CNIE) presents significant diagnostic and therapeutic challenges due to the absence of identifiable pathogens, leading to empirical antibiotic treatment and potentially worse clinical outcomes. This study aims to compare the clinical characteristics, surgical outcomes, and long-term prognosis of CNIE and culture-positive infective endocarditis (CPIE) patients.

**Methods:** A total of 698 patients who underwent surgery for infective endocarditis (IE) were included, with 154 (22.1%) classified as CNIE and 544 (77.9%) as CPIE. Baseline characteristics, preoperative risk factors, intraoperative variables, and postoperative outcomes were analyzed.

**Results:** CNIE patients exhibited significantly worse preoperative cardiac and renal function. Multivariate analysis identified female, perivalvular abscess and renal insufficiency are independent risk factors for early mortality in IE patients. Postoperatively, CNIE patients had a higher incidence of continuous renal replacement therapy (18% vs. 9%, p<0.01). Although long-term follow-up data showed no significant difference in the overall prognosis between CNIE and CPIE patients, subgroup analysis found that female CNIE patients showed worse long-term prognosis (HR=2.46 [1.16-5.23]; P=0.02).

**Conclusions:** Although the survival rates of CNIE and CPIE are similar, the prognosis of female CNIE patients is significantly worse.

**Clinical Perspective:**

- Culture-negative infective endocarditis (CNIE) patients who undergo surgery have favorable long-term outcomes comparable to culture-positive infective endocarditis (CPIE) patients, emphasizing the pivotal role of surgical intervention in infective endocarditis treatment.
- Female CNIE patients exhibit significantly worse surgical prognosis, highlighting the need for increased clinical attention and efforts to identify pathogens using other methods like tissue culture, PCR and mNGS.
- Although overall prognosis is similar, CNIE may indirectly worsen outcomes by impairing renal function, underscoring the importance of postoperative eGFR monitoring and protect kidney function.

## Introduction

The treatment of infective endocarditis (IE) remains a significant clinical challenge. With societal advancements and medical progress, the incidence of IE has been on the rise, yet its mortality rate remains persistently high ^1,2^. The type of causative pathogen plays a crucial role in determining treatment strategies and patient prognosis, making it a key focus for clinicians.

However, identifying the causative pathogen can be difficult due to factors such as prior antibiotic use, challenges in pathogen culture requiring specialized conditions, and advanced patient age. These factors often lead to culture-negative infective endocarditis (CNIE), complicating clinical decision-making ^3–5^. With the widespread use of antibiotics, the proportion of CNIE remains high, ranging from 5.2% to 31% in recent studies ^6,7^. This can result in treatment delays and the need for empirical antibiotic therapy, which will compromise the treatment efficacy.

Recently, only a few studies have analyzed the differences in surgical mortality in IE with or without identification of the pathogen, and they have obtained inconsistent results ^8,9^. Therefore, we conducted a study to explore whether the prognosis differs in these patients compared to those with CNIE.

## Methods

### Study Design

This study is a single-center retrospective cohort study that includes hospitalized patients with IE who underwent surgical treatment at Peking Union Medical College Hospital between January 2013 and December 2024. The study was approved by the Ethics Review Committee of Peking Union Medical College Hospital (I-24PJ0716). Given the retrospective nature of the study and the anonymization of patient identity information, the requirement for informed consent was waived by the ethics committee. In the inclusion selection, when analyzing the long-term prognosis of patients, 70 (10.0%) patients who were lost to follow-up were excluded (Supplementary Figure 1).

#### Definitions and Data Collection

IE was diagnosed based on the modified Duke criteria, then antibiotic treatment is first initiated by the infectious diseases team, followed by multidisciplinary team make evaluation for potential surgical intervention. CNIE was defined as cases in which at least two blood cultures (Different sampling sites, with sampling intervals greater than 12 hours) failed to identify a pathogen. Patients in whom the pathogen was identified by other diagnostic methods (like intraoperative tissue culture, PCR or mNGS) were classified as having culture-positive infective endocarditis (CPIE) for the purposes of analysis.

Patient data were collected from the hospital’s electronic medical record system, including demographic characteristics, clinical baseline information, laboratory and microbiological findings, echocardiographic imaging features, perioperative management data, and postoperative recovery indicators. Detailed definitions, units, and data types for each variable are provided in Supplementary Table 1. Follow-up data were collected by four cardiologists through medical record reviews and structured telephone interviews, achieving a follow-up rate of 90% (n = 628).

#### Statistical Analysis

All data analyses were performed using R Studio software (version 4.2.2; R Foundation for Statistical Computing, Vienna, Austria). Continuous variables were tested for normality by Shapiro-Wilk, and normally distributed data were expressed as mean ± standard deviation and compared using independent sample t test; non-normally distributed data were expressed as median (interquartile range, IQR) and analyzed using Mann-Whitney U test. Categorical variables were described as number of cases (percentages), and comparisons were performed using Pearson χ² test or Fisher’s exact probability method according to the sample size.

Variable screening was performed using stepwise regression (two-way elimination method) combined with clinical expert evaluation to ensure that the variables included in the model were both statistically significant and clinically relevant. The final screened predictive factors were included in the multivariate logistic regression analysis. For survival analysis, the Cox proportional hazards regression model was constructed to calculate the hazard ratio (HR) and its 95% confidence interval (CI). The cumulative survival rate was evaluated by plotting Kaplan-Meier survival curves, and the log-rank test was used for gender-stratified comparisons. All statistical tests were two-sided, and P < 0.05 was considered statistically significant. Mediation analysis was performed using structural equation modeling (SEM). Based on their correlation with CPIE (p < 0.1), three variables, eGFR, heart failure, and affect tricuspid valve, were selected as mediating variables. The model was resampled 5000 times using the bootstrap method to calculate bias-corrected confidence intervals.

To evaluate the robustness of the study results, we conducted sensitivity analysis on 1. Secondary endpoints (including sepsis, multiple organ failure (MOF), intra-aortic balloon pump (IABP)/extracorporeal membrane oxygenation (ECMO) use, and secondary thoracotomy). A multivariate logistic regression model was used to adjust confounding factors and analyze the association between CNIE and CPIE patients with secondary endpoints. 2. Patients were divided into two groups according to the time of surgery: before 2021 and after 2021 for stratified analysis to evaluate the impact of diagnosis and treatment strategies in different periods.

## Results

A total of 698 patients with IE who underwent surgical treatment were included in this study, with 544 cases (77.9%) in the CPIE and 154 cases (22.1%) in the CNIE.

### Baseline Characteristics

The baseline characteristics of both groups are summarized in Table 1 (overall and stratified). The median age of the overall population was 47 (34-57) years, with 476 (68%) being male, and there were no significant differences in baseline characteristics. Regarding comorbidities, there were no statistically significant differences in the prevalence of hypertension, diabetes, or chronic obstructive pulmonary disease (COPD) between the two groups. However, the proportion of CNIE patients with reduced LVEF (<50%) was significantly higher than that in CPIE group (10% vs. 3%, p<0.001), and the incidence of clinical heart failure was also higher (37% vs. 29%, p=0.047). In addition, CNIE had a higher incidence of moderate to severe renal impairment (eGFR <60 mL/min/1.73 m^2^: 21% vs 14%, p=0.02).

**Table 1.**
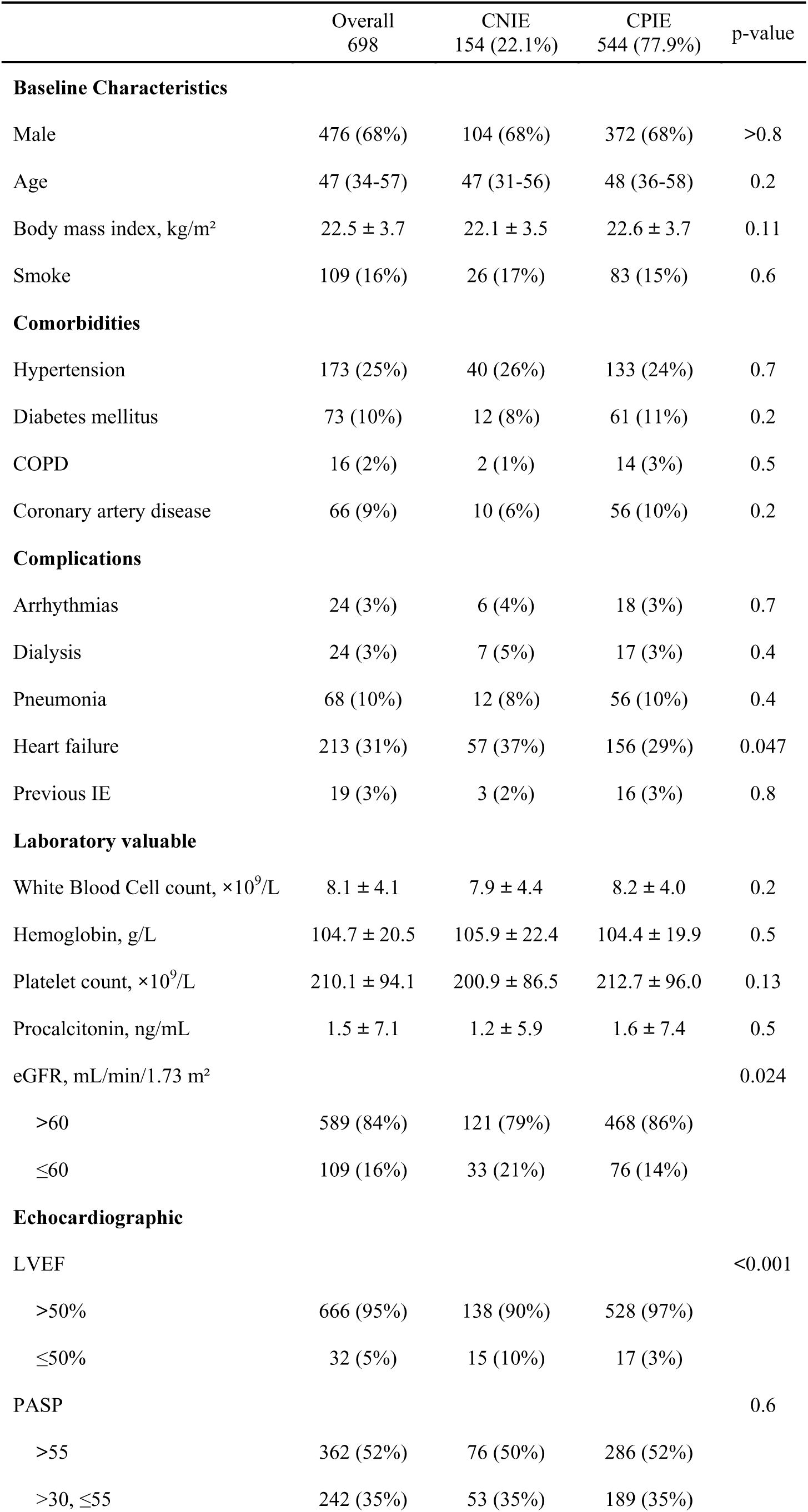
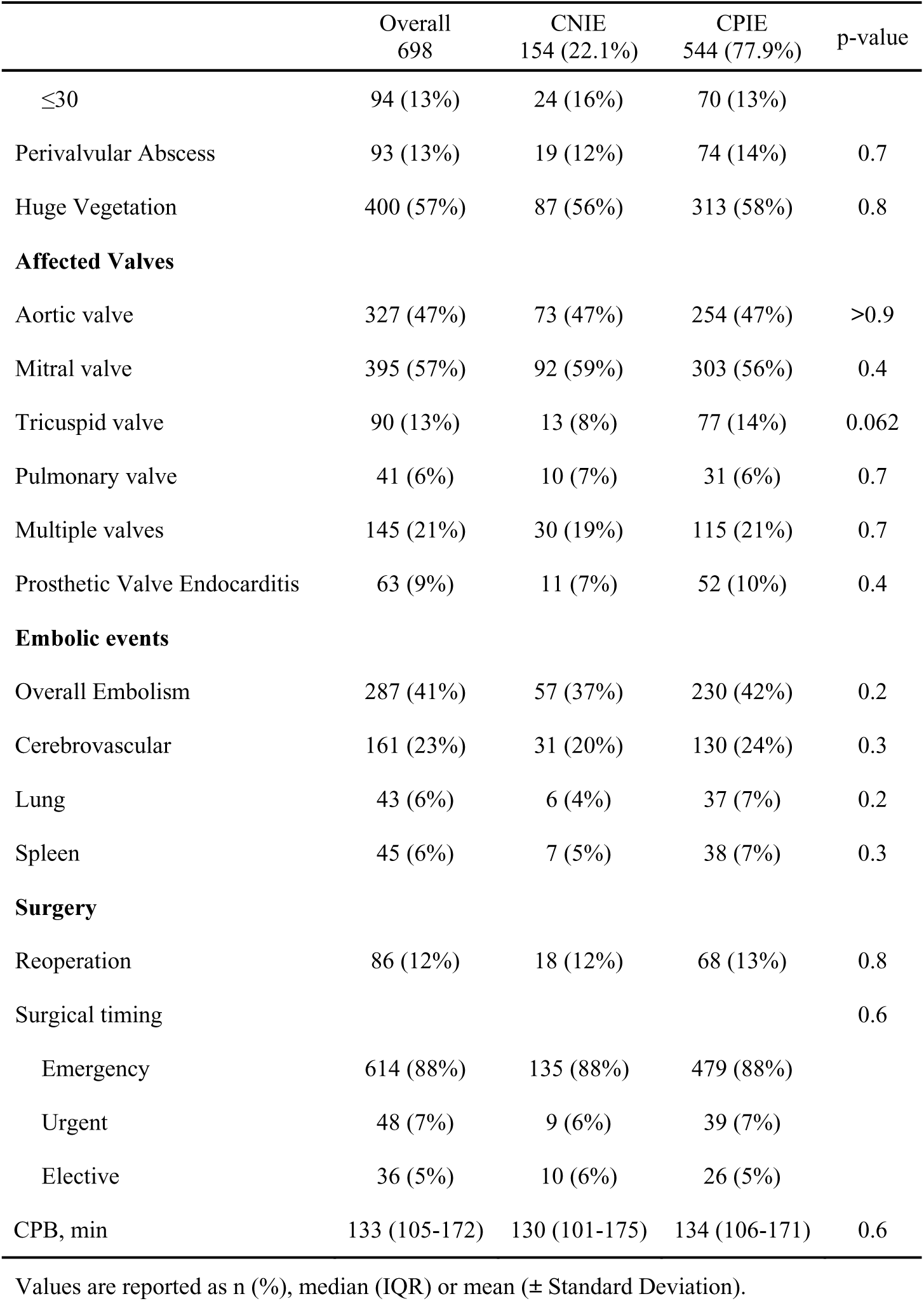
Baseline characteristics of the overall study population and stratified by CNIE in patients undergoing surgery for infective endocarditis.

|  | Overall<br>698 | CNIE<br>154 (22.1%) | CPIE<br>544 (77.9%) | p-value |
| --- | --- | --- | --- | --- |
| **Baseline Characteristics** |  |  |  |  |
| Male | 476 (68%) | 104 (68%) | 372 (68%) | >0.8 |
| Age | 47 (34-57) | 47 (31-56) | 48 (36-58) | 0.2 |
| Body mass index, kg/m<sup>2</sup> | 22.5 ± 3.7 | 22.1 ± 3.5 | 22.6 ± 3.7 | 0.11 |
| Smoke | 109 (16%) | 26 (17%) | 83 (15%) | 0.6 |
| **Comorbidities** |  |  |  |  |
| Hypertension | 173 (25%) | 40 (26%) | 133 (24%) | 0.7 |
| Diabetes mellitus | 73 (10%) | 12 (8%) | 61 (11%) | 0.2 |
| COPD | 16 (2%) | 2 (1%) | 14 (3%) | 0.5 |
| Coronary artery disease | 66 (9%) | 10 (6%) | 56 (10%) | 0.2 |
| **Complications** |  |  |  |  |
| Arrhythmias | 24 (3%) | 6 (4%) | 18 (3%) | 0.7 |
| Dialysis | 24 (3%) | 7 (5%) | 17 (3%) | 0.4 |
| Pneumonia | 68 (10%) | 12 (8%) | 56 (10%) | 0.4 |
| Heart failure | 213 (31%) | 57 (37%) | 156 (29%) | 0.047 |
| Previous IE | 19 (3%) | 3 (2%) | 16 (3%) | 0.8 |
| **Laboratory valuable** |  |  |  |  |
| White Blood Cell count, ×10<sup>9</sup>/L | 8.1 ± 4.1 | 7.9 ± 4.4 | 8.2 ± 4.0 | 0.2 |
| Hemoglobin, g/L | 104.7 ± 20.5 | 105.9 ± 22.4 | 104.4 ± 19.9 | 0.5 |
| Platelet count, ×10<sup>9</sup>/L | 210.1 ± 94.1 | 200.9 ± 86.5 | 212.7 ± 96.0 | 0.13 |
| Procalcitonin, ng/mL | 1.5 ± 7.1 | 1.2 ± 5.9 | 1.6 ± 7.4 | 0.5 |
| eGFR, mL/min/1.73 m<sup>2</sup> |  |  |  | 0.024 |
| >60 | 589 (84%) | 121 (79%) | 468 (86%) |  |
| ≤60 | 109 (16%) | 33 (21%) | 76 (14%) |  |
| **Echocardiographic** |  |  |  |  |
| LVEF |  |  |  | <0.001 |
| >50% | 666 (95%) | 138 (90%) | 528 (97%) |  |
| ≤50% | 32 (5%) | 15 (10%) | 17 (3%) |  |
| PASP |  |  |  | 0.6 |
| >55 | 362 (52%) | 76 (50%) | 286 (52%) |  |
| >30, ≤55 | 242 (35%) | 53 (35%) | 189 (35%) |  |
| ≤30 | 94 (13%) | 24 (16%) | 70 (13%) |  |
| Perivalvular Abscess | 93 (13%) | 19 (12%) | 74 (14%) | 0.7 |
| Huge Vegetation | 400 (57%) | 87 (56%) | 313 (58%) | 0.8 |
| **Affected Valves** |  |  |  |  |
| Aortic valve | 327 (47%) | 73 (47%) | 254 (47%) | >0.9 |
| Mitral valve | 395 (57%) | 92 (59%) | 303 (56%) | 0.4 |
| Tricuspid valve | 90 (13%) | 13 (8%) | 77 (14%) | 0.062 |
| Pulmonary valve | 41 (6%) | 10 (7%) | 31 (6%) | 0.7 |
| Multiple valves | 145 (21%) | 30 (19%) | 115 (21%) | 0.7 |
| Prosthetic Valve Endocarditis | 63 (9%) | 11 (7%) | 52 (10%) | 0.4 |
| **Embolic events** |  |  |  |  |
| Overall Embolism | 287 (41%) | 57 (37%) | 230 (42%) | 0.2 |
| Cerebrovascular | 161 (23%) | 31 (20%) | 130 (24%) | 0.3 |
| Lung | 43 (6%) | 6 (4%) | 37 (7%) | 0.2 |
| Spleen | 45 (6%) | 7 (5%) | 38 (7%) | 0.3 |
| **Surgery** |  |  |  |  |
| Reoperation | 86 (12%) | 18 (12%) | 68 (13%) | 0.8 |
| Surgical timing |  |  |  | 0.6 |
| Emergency | 614 (88%) | 135 (88%) | 479 (88%) |  |
| Urgent | 48 (7%) | 9 (6%) | 39 (7%) |  |
| Elective | 36 (5%) | 10 (6%) | 26 (5%) |  |
| CPB, min | 133 (105-172) | 130 (101-175) | 134 (106-171) | 0.6 |

Values are reported as n (%), median (IQR) or mean (± Standard Deviation).

In echocardiographic results, there were no significant differences in the incidence of paravalvular complications and the size of vegetation between the two groups. IE most commonly affected the left heart (83%), while the proportion of tricuspid valve involvement in CNIE showed an increased trend (14% vs. 8%, p=0.066), but it did not reach statistical significance. The most common preoperative embolic event was ischemic stroke (n=161, 23%), with no difference between the two groups (P=0.3); the incidence of splenic embolism and pulmonary embolism was similar (n=43, n=45, respectively). In terms of surgical intervention, the two groups were comparable in the timing of surgery (emergency surgery ratio: 6% vs. 5%, p=0.6).

### Postoperative Complications

The postoperative complications analysis showed in Table 2, which revealed that the need for continuous renal replacement therapy (CRRT) after surgery was significantly higher in the CNIE group (18% vs. 9%, p < 0.01). Although the incidence of postoperative low cardiac output syndrome (LCOS) was higher in the CNIE group compared to the control group (12% vs. 8%), the difference did not reach statistical significance (p = 0.12). Similarly, there were no statistically significant differences between the two groups in the use of circulatory assist devices, like IABP, ECMO, and permanent pacemaker implantation (all p > 0.05).

**Table 2.** Comparison of postoperative complications among the overall patients and each subgroup.

|  | Overall<br>698 | CNIE<br>154 (22.1%) | CPIE<br>544 (77.9%) | p-value |
| --- | --- | --- | --- | --- |
| CNS | 18 (3%) | 1 (1%) | 17 (3%) | 0.14 |
| Long ventilation | 163 (23%) | 38 (25%) | 125 (23%) | 0.7 |
| Pneumonia | 80 (11%) | 17 (11%) | 63 (12%) | 0.9 |
| Atrial fibrillation | 101 (14%) | 18 (12%) | 83 (15%) | 0.3 |
| Atrioventricular block | 23 (3%) | 3 (2%) | 20 (4%) | 0.3 |
| Re-thoracotomy | 42 (6%) | 11 (7%) | 31 (6%) | 0.5 |
| Low Cardiac Output Syndrome | 60 (9%) | 18 (12%) | 42 (8%) | 0.12 |
| Multiple Organ Failure | 24 (3%) | 7 (5%) | 17 (3%) | 0.4 |
| Septic shock | 31 (4%) | 6 (4%) | 25 (5%) | 0.7 |
| Machine Auxiliary Device |  |  |  |  |
| IABP | 16 (2%) | 7 (5%) | 9 (2%) | 0.060 |
| ECMO | 14 (2%) | 3 (2%) | 11 (2%) | >0.9 |
| Pacemaker | 17 (2%) | 4 (3%) | 13 (2%) | 0.8 |
| CRRT | 77 (11%) | 28 (18%) | 49 (9%) | 0.001 |
| Treatment Time |  |  |  |  |
| ICU time, days | 2 (1-5) | 2 (1-5) | 2 (1-5) | 0.4 |
| Hospital time, days | 10 (7-14) | 11 (8-16) | 10 (7-14) | 0.13 |
| In-hospital mortality | 13 (2%) | 2 (1%) | 11 (2%) | 0.7 |

Values are reported as n (%), median (IQR) or mean ( ± Standard Deviation).

### Ending and Fellow-up

A total of 13 patients (2%) died during hospitalization. After a median follow-up duration of 4.7 years (IQR 2.2–7.1), survival analysis demonstrated comparable outcomes between CNIE and CPIE patients at 1-month (97% vs 97%; HR=0.97 [0.32-2.92], P=0.96) and 1-year survival rates (90% vs 92%; HR=1.18 [0.63-2.21], P=0.6) (Supplementary Table 2 and Supplementary Figure 2). Multivariate analysis identified male (OR=0.32 [0.11, 0.89]; P=0.03), perivalvular complications (HR=4.07 [1.16, 14.1]; P=0.03) and eGFR (HR=4.51 [1.47, 13.7]; P<0.01) as significant factors associated with early mortality (Table 3). Figure 1 shows that there was no significant difference in the cumulative mortality rate between CPIE and CNIE patients (89% vs 86%; HR=1.21 [0.72-2.03]; P=0.47). However, gender-stratified multivariate analysis showed that the clinical prognosis of female CNIE patients was significantly worse than that of CPIE patients (HR=2.46 [1.16-5.23]; P=0.02) (Figure 2).

**Figure 1.**
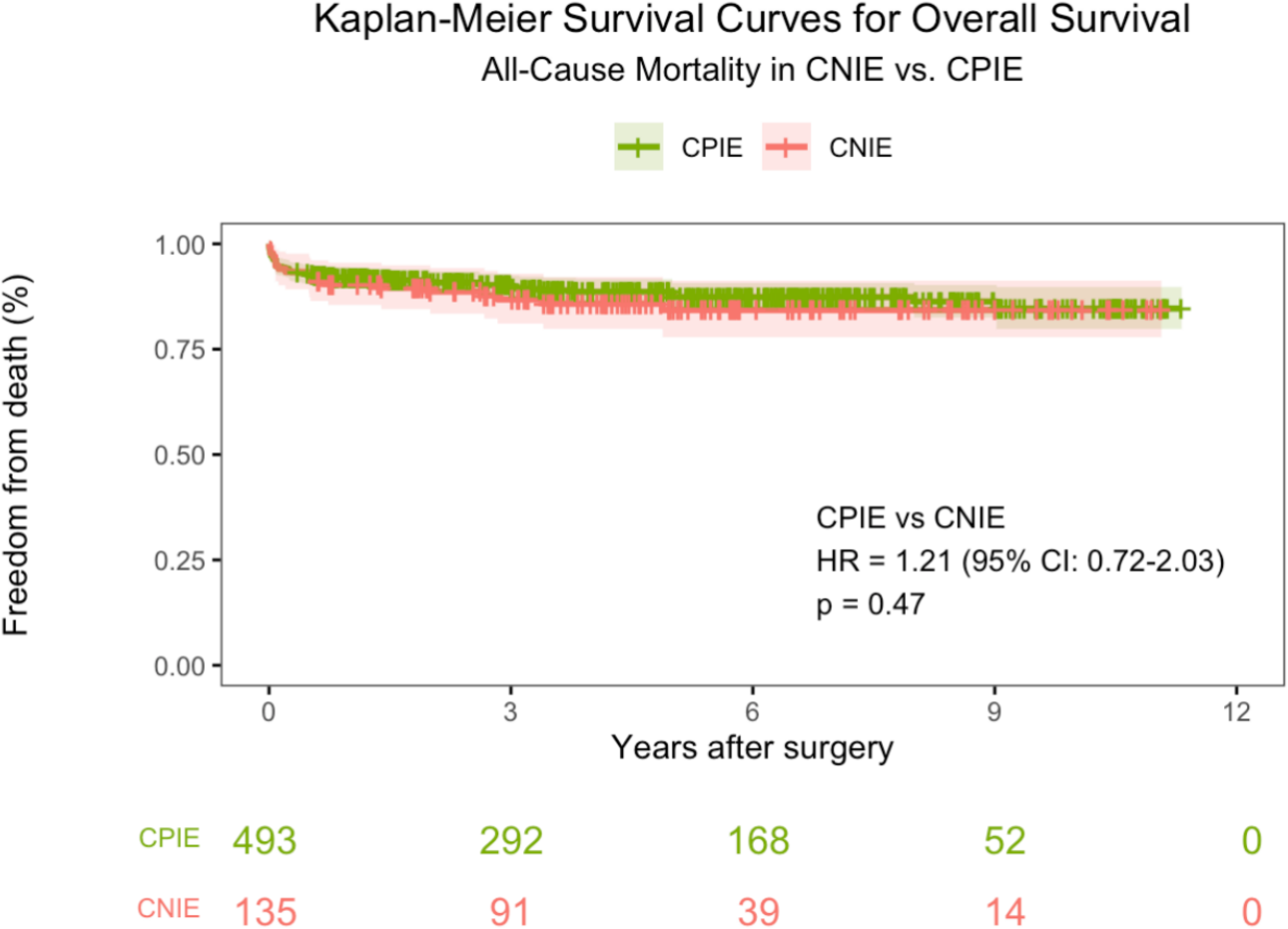
Kaplan–Meier Survival Curves for overall survival in patients between CNIE and CPIE.

**Figure 2.**
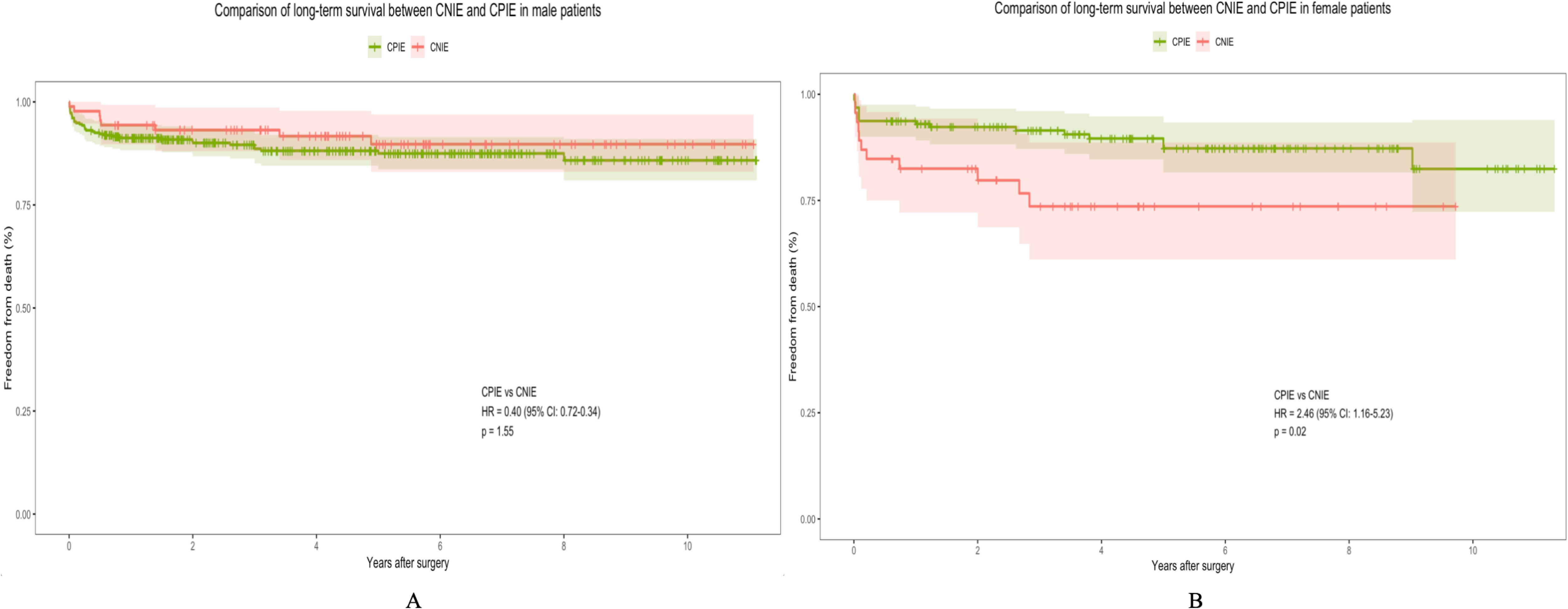
The Kaplan–Meier method was used to analyze differences in survival rates between patients with CPIE (green curve) and with CNIE (red curve). Shaded areas indicate the 95% confidence intervals. A: Survival status in female patients; B: Survival status in male patients.

**Table 3.** Univariable and multivariable analyses of early mortality in 698 patients undergoing surgery for infective endocarditis.

|  | Univariable analysis |  | Multivariable analysis |  |
| --- | --- | --- | --- | --- |
|  | OR [95% CI] | p-value | OR [95% CI] | p-value |
| Age | 1.03 [0.99, 1.06] | 0.11 | 1.02 [0.98, 1.06] | 0.25 |
| Male | 0.42 [0.17, 1.07] | 0.07 | 0.32 [0.11, 0.89] | 0.03 |
| eGFR≤60mL/min/1.73 m<sup>2</sup> | 5.47 [2.12, 14.0] | <0.01 | 4.51 [1.47, 13.7] | <0.01 |
| Diabetes Mellitus | 3.21 [1.01, 8.71] | 0.03 | 1.87 [0.50, 6.04] | 0.33 |
| Culture-Positive Infective Endocarditis | 0.97 [0.27, 2.73] | 0.96 | 0.83 [0.20, 2.74] | 0.77 |
| Reoperation | 4.35 [1.57, 11.2] | <0.01 | 3.75 [0.23, 25.1] | 0.30 |
| LVEF≤50% | 1.15 [0.06, 5.90] | 0.89 | 1.13 [0.06, 7.30] | 0.92 |
| Perivalvular Abscess | 5.18 [1.95, 13.2] | <0.01 | 4.07 [1.16, 14.1] | 0.03 |
| CNS | 1.53 [0.53, 3.95] | 0.40 | 2.70 [0.86, 8.03] | 0.09 |
| Native Valve Endocarditis | 0.20 [0.07, 0.58] | <0.01 | 1.48 [0.08, 12.9] | 0.75 |

CNS: Disease involve the central nervous system;

Through mediation analysis, it was found that CPIE itself did not have a significant effect on patient survival (OR = 0.993 [0.929, 1.062], p = 0.827). However, the total indirect effect was significant, with the indirect path through eGFR influencing the risk of death (OR = 0.988 [0.975, 1.002], p = 0.068) (Supplementary Table 3).

Sensitivity analysis further showed that there was no statistical difference in the risk of secondary outcomes between CNIE and CPIE (OR=1.29 [0.69-2.34]; P=0.42), and the lower eGFR and paravalvular abscess were still independent risk factors for secondary outcomes (Supplementary Table 4). In addition, stratified analysis by surgical year (before or after 2021) also found no significant effect of CNIE on prognosis (before 2021: HR=1.56 [CI 0.86-2.83], P=0.15; after 2021: HR=0.57 [CI 0.17-1.89], P=0.36) (Supplementary Figure 3).

## Discussion

This study examined the long-term outcomes of 698 IE patients, comparing the clinical characteristics of CNIE and CPIE. CNIE patients had lower ejection fraction, and they had lower preoperative eGFR, contributing to a higher incidence of postoperative renal failure. Mortality analysis showed that elderly, perivalvular complications, lower eGFR and CNS were risk factors for IE patients. Although the overall analysis showed no significant difference in survival between CNIE and CPIE patients, gender-stratified analysis found that the prognosis of female CNIE patients was significantly worse than that of the CPIE group (HR=0.41 [0.19-0.86]; P=0.02), while there was no difference between the two groups for male patients.

The inability to identify the causative pathogen in IE patients presents a significant clinical challenge. CNIE often require empirical antibiotic therapy due to the lack of precise targeted treatment, which may result in insufficient coverage or improper dosing, thereby increasing in-hospital mortality risk ^10^. The most common reason for CNIE is prior antibiotic use ^11^, while true CNIE is typically associated with fastidious organisms that are difficult to detect using conventional methods, such as the HACEK group, Coxiella burnetii (Q fever), Bartonella species, and so on ^12–14^. Although some studies suggest that CNIE may have a milder clinical presentation than CPIE ^15^, our findings indicate that among patients requiring surgical intervention, CNIE exhibit worse preoperative conditions. These findings align with Zamorano et al.’s study, which demonstrated that CNIE are more likely to present advanced heart failure and significant valvular destruction ^16^.

CNIE tend to have lower glomerular filtration rates, which may be attributed to multiple factors. Firstly, these patients often receive prolonged empirical antibiotic therapy to manage unexplained fever, and some antibiotics have nephrotoxic effects that can further exacerbate kidney damage ^17^. Additionally, in certain cases, underlying autoimmune diseases may contribute to the formation of vegetations and persistent fever, with immune complex deposition and vasculitic glomerulonephritis further impairing renal function. Hemodynamic disturbances caused by heart failure may also reduce renal perfusion, worsening kidney dysfunction. Studies have shown that both cerebral embolism and renal impairment are significant predictors of poor outcomes ^18^. CNIE patients often have renal impairment, which not only significantly increases the demand for CRRT, but also may worsen the patient’s prognosis and thus reduce patient’s quality of life.

Multivariate analysis showed that age, paravalvular complications, eGFR grade, and neurological complications were independent risk factors for death in surgical patients. Previous studies have found that independent risk factors affecting the prognosis of IE patients include renal insufficiency, impaired cardiac function, large vegetations, neurological complications, and paravalvular complications ^19,20^. Marouane et al. also showed that the in-hospital mortality rate of patients over 80 years old was significantly increased ^21^.

Regarding the factors affecting the postoperative prognosis of patients with CNIE, there are still differences in the conclusions of existing studies. Toshifumi et al. proposed that CNIE, postoperative heart failure and prosthetic valve infection are independent risk factors for long-term prognosis ^8^, while Di et al.’s study showed that postoperative mortality was not significantly correlated with blood culture results ^9^. This difference may be related to factors such as study population characteristics, treatment strategies (such as empirical use of antibiotics) or follow-up duration. It is worth noting that although mainstream IE prognostic scoring systems (such as PALSUSE and ENDOFORS) do not include CNIE as a core variable ^22–24^, recent evidence shows that integrating blood culture status can improve the predictive power of the model (AUC increased from 0.684 to 0.71, p=0.439), suggesting its potential prognostic value ^25^. This study demonstrated no statistically significant difference in long-term prognosis or the incidence of major complications between CNIE and CPIE patients who underwent surgical treatment. These findings are consistent with those reported in the ESC-EORP EURO-ENDO registry^7^, further confirms the effectiveness of surgical intervention in infective endocarditis with unidentified pathogen type. Although the long-term survival rates between CNIE and CPIE patients did not differ significantly, mediation analysis revealed a potential underlying mechanism. CNIE was associated with an increased risk of mortality by accelerating the deterioration of renal function, as evidenced by a progressive decline in eGFR, which suggests that, regular monitoring of dynamic eGFR changes is essential, early identification of renal function decline should be prioritized, and proactive renal protection strategies should be implemented.

In clinical studies of infective endocarditis (IE), the impact of gender differences on prognosis has always been controversial ^26–28^. This study found through stratified analysis that the risk of death in female patients with infective endocarditis (CNIE) was significantly increased. This gender difference may be related to the fact that female patients are more likely to have delayed diagnosis and treatment ^29^. On the other hand, the relatively smaller cardiac anatomical structure of women increases the difficulty of surgical operation, thus affecting prognosis ^30^. Based on the above research results, it is recommended to use molecular diagnostic techniques (such as broad-spectrum PCR and mNGS) for CNIE patients (especially women) who require surgical intervention to accurately identify pathogens and optimize perioperative antimicrobial treatment regimens, thereby improving long-term prognosis.

## Data Availability

The datasets generated and/or analyzed during the current study are not publicly available due to patient privacy and institutional regulations but are available from the corresponding author on reasonable request and with permission from Peking Union Medical College Hospital.

## Legends

Supplementary Figure 1. Flow chart

Supplementary Figure 2. The Kaplan-Meier method was used to analyze the difference in survival rate between patients with positive blood culture (green curve) and negative blood culture (red curve). The shaded area represents the 95% confidence interval. A. the survival status during the 1-month follow-up period; B. the survival status during the 1-year follow-up period.

Supplementary Figure 3. Comparison of Kaplan-Meier survival curves of patients with CPIE (green curve) and CNIE (red curve) by year of surgery. A. Patients who underwent surgery before 2021 (n=260). B. Patients who underwent surgery after 2021 (n=368).

Abbreviated Legend. Kaplan–Meier Survival Curves for overall survival in patients between CNIE and CPIE.

## Glossary of Abbreviations

CNIE: Culture-negative infective endocarditis
CPIE: Culture-positive infective endocarditis
IE: Infective endocarditis
eGFR: Estimated glomerular filtration rate
LVEF: Left ventricular ejection fraction
CRRT: Continuous renal replacement therapy
LCOS: Low cardiac output syndrome
IABP: Intra-aortic balloon pump
ECMO: Extracorporeal membrane oxygenation
PCR: Polymerase chain reaction
mNGS: Metagenomic next-generation sequencing
MOF: Multiple organ failure
CNS: Central nervous system
COPD: Chronic obstructive pulmonary disease
TV: Tricuspid valve
SEM: Structural equation modeling

